# How vaccination and contact isolation might interact to suppress transmission of Covid-19: a DCM study

**DOI:** 10.1101/2021.01.03.20248972

**Authors:** Karl J. Friston, Anthony Costello, Guillaume Flandin, Adeel Razi

## Abstract

This report describes a dynamic causal model that could be used to address questions about the rollout and efficacy of vaccines in the United Kingdom. For example, is suppression of community transmission a realistic aspiration? And, if not, what kind of endemic equilibrium might be achieved? What percentage of the population needs to be vaccinated? And over what timescale? It focuses on the synergies among (i) *vaccination*, (ii) the supported *isolation of contacts* of confirmed cases and (iii) *restrictions on contact rates* (i.e., lockdown and social distancing). To model these mitigations, we used a dynamic causal model that embeds an epidemiological model into agent-based behavioural model. The model structure and parameters were optimised to best explain responses—to the first and subsequent waves—enabling predictions over the forthcoming year under counterfactual scenarios. Illustrative analyses suggest that the full potential of vaccination is realised by increasing the efficacy of contact tracing: for example, under idealised (best case) assumptions—of an effective vaccine and efficient isolation of infected pre-symptomatic cases— suppression of community transmission would require 50% herd immunity by vaccinating 22% by the end of 2021; i.e., 15 million people or about 50,000 per day. With no change in the isolation of contacts, 36% would require vaccination, i.e., 25 million people. These figures should not be read as estimates of the actual number of people requiring vaccination; however, they illustrate the potential of this kind of model to quantify interactions among public health interventions. We anticipate using this model in a few months—to estimate the average effectiveness of vaccines when more data become available.

## Introduction

With the announcement of regulated vaccines and an imminent programme of vaccination, there are several pressing questions about the rate and extent of vaccination required to contain and possibly suppress the spread of SARS-CoV-2 (Saad-Roy, Wagner et al. 2020)^1^. Established quantitative criteria can provide guidance on the rollout and extent of requisite vaccination programs^2^. For example, it is generally considered that a herd immunity of 67% of population will be sufficient to suppress community transmission of SARS-CoV-2. These kinds of estimates usually assume an idealised epidemiology, in which every member of the population is exchangeable, and the reproduction ratio is fixed, here, R_0_ = 3. However, heterogeneity in exposure, susceptibility and transmission—coupled with fluctuating reproduction ratios—may mean that the proportion of the population requiring vaccination may be less than 67% (Fontanet and Cauchemez 2020, Gomes, Corder et al. 2020, Lourenco, Pinotti et al. 2020). Furthermore, pre-existing immunity (Le Bert, Tan et al. 2020, Ng, Faulkner et al. 2020) may further reduce the number of people who need to be vaccinated. In what follows, we use dynamic causal modelling to assimilate information from past responses to the outbreak to predict what one might expect under different vaccination scenarios in the future. This offers a slightly more nuanced treatment that considers vaccination in the context of heterogeneity, ongoing restrictions and public health measures.

The purpose of this technical note is to describe the model in question and illustrate its application to data at the time of writing (6-12-2021). Its purpose is to provide a coarse-grained way of assessing the average effectiveness of public health measures and, in particular, how they contextualise each other. The analyses that follow are therefore presented purely for illustrative purposes. We anticipate using this model to quantify the effectiveness of vaccination in a few months when appropriate data become available.

Dynamic causal modelling may play a useful role in this setting because it features a state-space model of both *epidemiological and behavioural* responses to infection at a population level. Furthermore, some of these responses can be characterised as interventions, enabling model fitting—based upon empirical data from the past—to inform the structure and parameters that underwrite viral spread in the future (Friston, Parr et al. 2020, Friston, Parr et al. 2020). Here, we use a dynamic causal model to generate posterior predictive densities over some key outcomes (e.g., fatality rates, confirmed cases and vaccination rates). This dynamic causal model (DCM) has been progressively optimised using Bayesian model comparison during the course of the epidemic—to ensure that it has the right balance of expressivity (i.e., model complexity) and predictive accuracy. The details of this model can be found in (Friston, Flandin et al. 2020), along with (i) the Master equation that underwrites the generative model, (ii) the prior over model parameters (and their sources) and (iii) the variational scheme used for model inversion and comparison.

For people not familiar with dynamic causal modelling (DCM), it can be regarded as an analytic version of a stochastic agent-based model, where the sample densities are replaced by a mean field approximation. This allows one to model the ensemble density dynamics analytically. In turn, this affords an efficient way of fitting the model to observed time series (i.e., data assimilation), while, at the same time, quantifying uncertainty about model parameters. Most importantly, it allows one to score each model in terms of (a variational bound on) model evidence; thereby enabling model comparison or structure learning. A model with a high evidence (a.k.a. marginal likelihood) has greater predictive validity in the sense that the likelihood of new data, under the posterior predictive density will, on average, be greater. Crucially, model evidence is a function of the data, which means the predictive validity of the model with the greatest evidence increases with the available data. We use this feature of DCM to licence its application to the scenario modelling at hand.

Our focus is on the dynamics of vaccination and interactions with other interventions; in particular, the adaptive *restriction of contact rates* through social distancing, lockdowns, and travel restrictions. In the current model, these mechanisms are lumped together in terms of a single probability that any member of the population will move from a low (e.g., home) to a high risk (e.g., work) location on any given day. This contact rate is itself a function of the prevalence of infection. This function models things like ‘tier systems’ and ‘circuit breakers’—as implemented as a governmental level—but, ultimately, driven by (measures of) the prevalence of infection and its fluctuations (e.g., the reproduction ratio or R-number). In brief, contact rates are a non-linear threshold function of a linear mixture of various prevalence levels (including the number of people who are currently infected and seroprevalence). Please see (Friston, Flandin et al. 2020) for details. The second important mitigating mechanism is the supported *isolation of contacts*; namely, reducing viral transmission by minimising the contact rates with people who are infected but have yet to become infectious. Of all the mitigating mechanisms, this is the most efficient; in the sense that it is the most specific. In other words, it targets the small proportion of the population that can transmit the virus to others. This should be contrasted with restricting contact rates through social distancing: this mechanism applies to everyone and is more costly to implement at several levels.

For example, consider heterogeneity of transmission due to overdispersion (Lloyd-Smith, Schreiber et al. 2005, Endo, Abbott et al. 2020, Gomes, Corder et al. 2020). This is sometimes cast in terms of superspreading; namely, when the few infect the many. Assume, for example, that only 20% of the population can transmit the virus to someone else. If one were able to identify superspreaders, then it would only be necessary to vaccinate—or isolate—20% of the population to eliminate community transmission. However, there is no way of knowing who is likely to infect someone else in advance. Or is there? This, on one view, is the rationale for contact tracing (critically, backward contact tracing or ‘cluster busting’). In other words, one can identify a subpopulation with an enriched probability that it contains people who are likely to infect others in the near future. This is why contact tracing and supported isolation has played a key role in containing the virus in many countries with established public health measures. One of our key questions was whether vaccination would enable efficient contact tracing in the United Kingdom by reducing the prevalence of infection.

To address this question, we simulated different vaccination rates with and without enhanced contact tracing to quantify their interaction. This was a simulated under context-sensitive restrictions on contact rates that best explain population behaviour to date.

### Dynamic causal modelling

To model the impact of vaccination, we introduced a new rate parameter determining the probability of moving from a susceptible (non-infected) state to a state of *seronegative* immunity, when visiting hospital (or an equivalent location). Vaccination was restricted to clinical settings because, in the model, these are frequented by clinically susceptible people and healthcare workers, who we presumed would be the first people to receive the vaccine. In the model, vaccination is taken to be hundred percent effective in precluding infection and thereby transmission. A single *effective vaccination* will therefore generate multiple recorded vaccinations, depending upon the number of vaccinations required for any given person, and their average *effectiveness*. We hope to estimate vaccine effectiveness, when the number of vaccinations administered over the next few months becomes available (using priors from the appropriate Phase 3 clinical trials).

In this DCM, there are two kinds of protective (humoral) immunity labelled *seropositive* and *seronegative*. The loss of seropositivity is modelled as a movement from seropositive to seronegative, with an estimated time constant of about 200 days. One then loses protective immunity by moving from a *seronegative* state to a *susceptible* state with a longer time constant (here, 2048 days)^3^. We modelled vaccination in this way because seroconversion induced by viral infection drives reduced contact rates—and we did not want to conflate vaccination induced seroconversion with seroprevalence due to infection. Furthermore, we anticipate that surveillance studies of seropositivity will deal separately with people who have and have not been vaccinated. Note that the seronegative state does not imply the absence of antibodies, rather the absence of antibodies following infection.

One can regard the seronegative state as an enduring state of immunity that does not entail seroconversion due to infection: i.e., an immune state due to pre-existing immunity (Le Bert, Tan et al. 2020, Ng, Faulkner et al. 2020), mediated by T-cells (Grifoni, Weiskopf et al. 2020, Le Bert, Tan et al. 2020, Seo, Giles et al. 2020) or vaccination (Fontanet and Cauchemez 2020, Houlihan, Vora et al. 2020, Saad-Roy, Wagner et al. 2020). This construction allows for the loss of detectable IgG antibodies (seropositivity) over months, while modelling a slower loss of protective (seronegative) immunity over years. Effective vaccination is assumed to induce an immunity of the second kind.

As previously, contact tracing is modelled with a similar rate parameter determining the probability of going into self-isolation when infected but pre-symptomatic. This models people’s response when advised by a family member, public health worker or contact tracer that they may be infected. The efficiency of contact tracing corresponds to this rate parameter. Its estimated level is about 3 to 4% based upon our past responses (see also ^4^). An efficient contact tracing is considered to be about 24%. This is based upon an idealised ability to detect 30% of new cases every day and support 80% of their contacts in self-isolation.

The parameter estimates above were based upon Bayesian model inversion using 12 sources of empirical data, shown in Figure 1. These include daily confirmed cases by specimen date, deaths with a positive PCR test within 28 days by date of death, hospital admissions, patients requiring mechanical ventilation, certified deaths, symptoms, mobility data provided by the Department of Transport, and so on. Collectively, these time series afford multiple constraints on the posterior probability of the unknown model parameters that are, universally, nonnegative rate or time constants^5^. The form and parameters of this model have been optimised using data from the first and secondary waves. This is important because assimilating data from the secondary wave mandated the inclusion of monotonic and seasonal fluctuations that, in principle, lend the model a greater predictive validity—in terms of long-term forecasting over months and years^6^. Equipped with these posterior expectations, one can roll out the probability distribution over various states into the future, under different sets of parameters.

**Figure 1:**
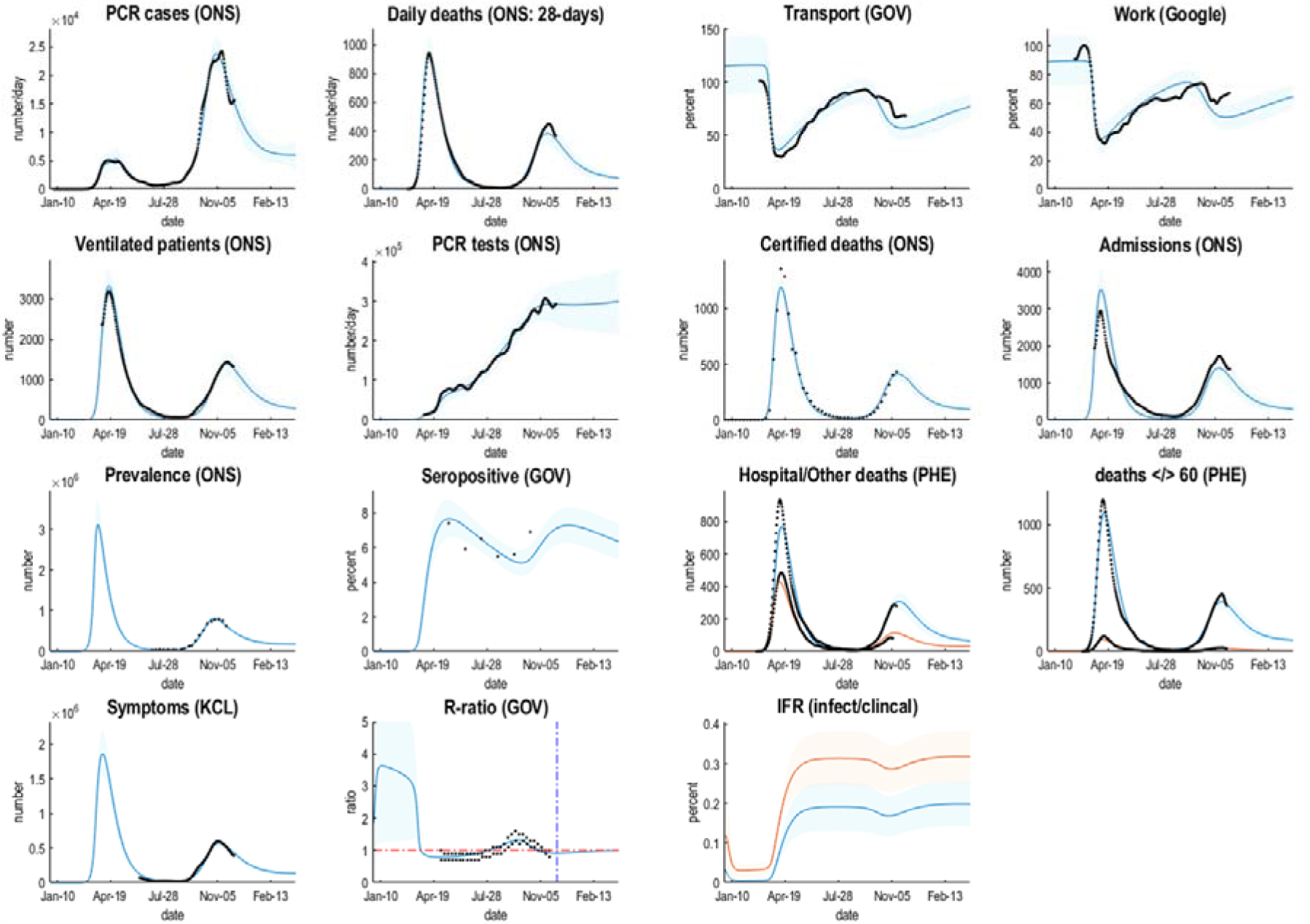
Posterior predictions of various outcomes, ranging from confirmed cases through to the reproduction ratio from the following sources: https://coronavirus.data.gov.uk; https://www.ons.gov.uk/peoplepopulationandcommunity/healthandsocialcare/conditionsanddiseases/datasets/coronaviruscovid19infectionsurveydata; https://covid.joinzoe.com/data#levels-over-time; https://www.gov.uk/guidance/the-r-number-in-the-uk#contents; https://www.gov.uk/government/statistics/transport-use-during-the-coronavirus-covid-19-pandemic; https://www.google.com/covid19/mobility/. The key thing to take from these results is the ability of the model to provide a fairly accurate account of multiple aspects of the epidemic; ensuring an internal consistency to the explanations furnished by the dynamic causal model. The coloured lines correspond to posterior expectations, while the shaded areas correspond to 90% credible intervals. The black dots are the empirical timeseries used to estimate the model parameters and their posterior uncertainty. The final panel provides estimates of infection fatality ratios (IFR) based upon the cumulative deaths and estimated infections or symptomatic cases (i.e., a clinical COVID-19 case). These ratios (0.18% and 0.32%) are substantially less than usually reported (Birrell, Blake et al. 2020, Russell, Hellewell et al. 2020, Streeck, Schulte et al. 2020, Verity, Okell et al. 2020), which may speak to an underestimation of prevalence based on serology or PCR testing. However, they are quantitatively consistent with surveys of ‘sentinel’ populations; for example, “In Manaus, the IFRs were 0.17% and 0.28%, considering PCR confirmed COVID19 deaths and probable COVID-19 deaths based on syndromic identification.” (Buss, Prete et al. 2020). The DCM estimates of IFR increase to about 0.5% when using the number of people infected, as opposed to the number of infections. These estimates suggest that SARS-CoV-2 is several times more virulent than seasonal influenza, when averaged across all ages.

To model different levels of vaccination, we increased the vaccination rate from 0% to 100% in 20% steps. By construction, a 100% vaccination rate (when visiting a hospital or healthcare centre) corresponds to a peak vaccination rate of about 200,000 per day, dropping to 50,000 per day after a month or two. If sustained over a year, this would ensure that about 28 million people would have received a course of vaccination by 2022. This (maximal) vaccination rate is comfortably within current public health capabilities. For example, current reports suggest that the NHS intends to use football stadiums, town halls and conference buildings in England to inoculate at least 2,000 people per centre each day. These new facilities will supplement the 1,560 community-based vaccination centres run by general practitioners, that can dispense between 200 to 500 vaccinations a day.

We did not consider higher rates of vaccination because, in this model, being vaccinated confers 100% seronegative immunity. To the extent that vaccination is only partially effective, the vaccination rates need to be adjusted accordingly. For example, if vaccination is only 50% effective, then the actual vaccination rates should be doubled. Similarly, if vaccinations are to be dispensed twice, the number of vaccinations should be doubled. Modelling vaccination as an *effective vaccination* means the efficacy of the vaccine does not influence the requisite herd immunity or number of people who need to receive an *effective vaccination*.

## Results

We increased the vaccination rate from zero to 100% under two levels of contact tracing (4% and 24%). Figure 2 shows the posterior predictive densities over mortality and mobility at current levels of contact tracing (4%). The corresponding trajectories of latent states are shown in Figure 3. To measure the impact of vaccination, we assessed cumulative mortality until the end of 2022. This was supplemented with one of many proxies for economic cost; namely, the percentage change in mobility based upon predictions of car use, relative to pre-COVID levels. We also looked at the reproduction ratio under different vaccination rates.

**Figure 2:**
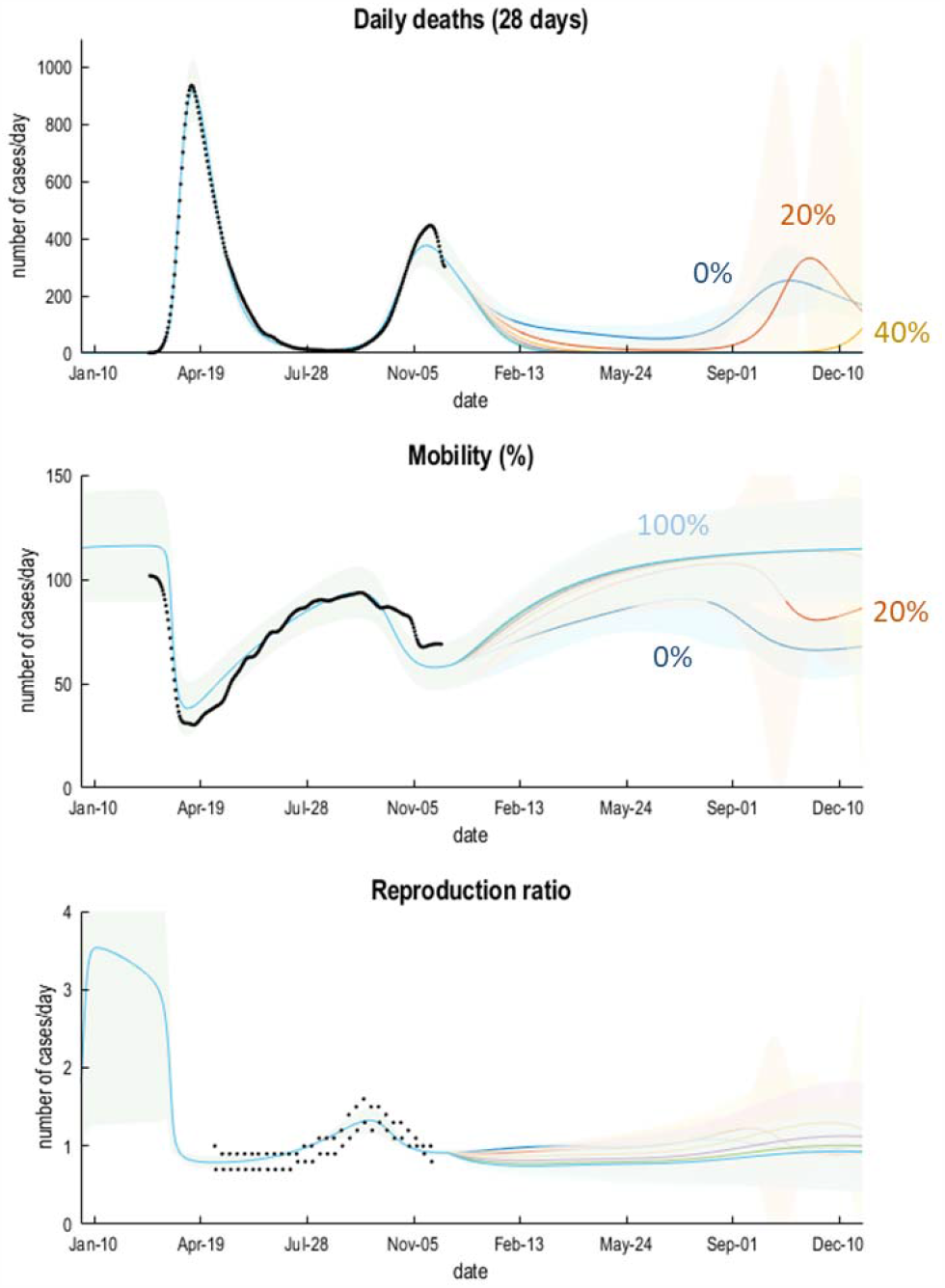
posterior predictions of fatalities, mobility and viral replication as scored by the reproduction ratio or R-number. The coloured lines correspond to the posterior expectations under six vaccination rates starting from 8 December 2021. The shaded areas correspond to 90% Bayesian credible intervals. The black dots correspond to empirical data sourced according to Figure 1.

**Figure 3:**
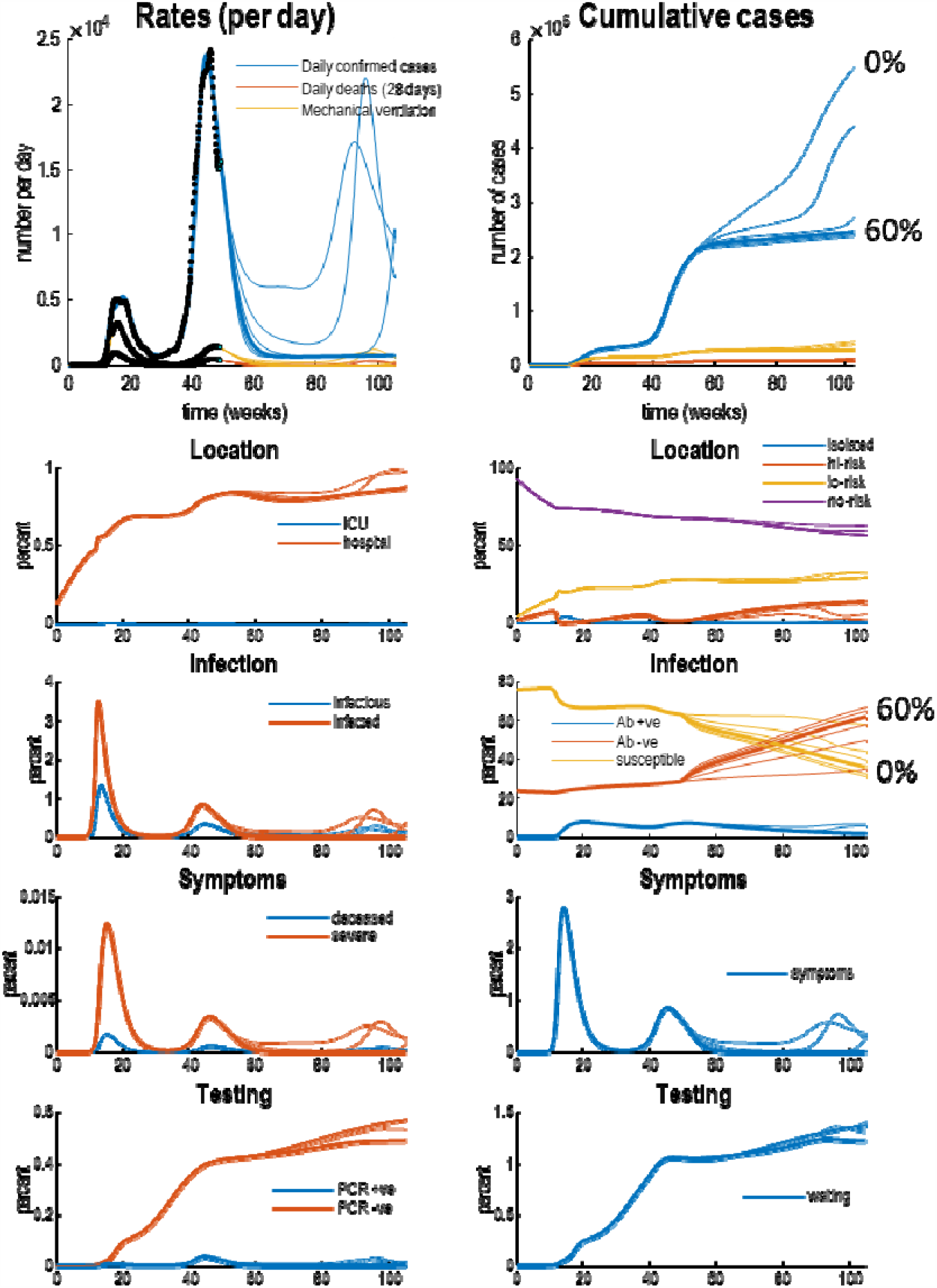
posterior predictions and underlying latent states. The upper panels show posterior expectations (coloured lines) and data (dots). The lower panels show the underlying posterior estimates of latent states generating these data. Each set of coloured lines corresponds to 6 levels of vaccination rates (increasing from 0% to 100% in steps of 20%). The thicker lines indicate trajectories under a 60% vaccination rate. Crucially, vaccination rates of 60% or more suppress viral transmission. The slow accumulation of confirmed cases (blue lines in the upper two panels) is due to a non-zero false positive rate under sustained testing, that includes false-positives post-vaccination. Recall that the seronegative immune state (Ab -ve) accumulates people who have been effectively vaccinated, while seropositive (Ab +ve) corresponds to those people who have been infected naturally. Please see (Friston, Flandin et al. 2020) for a full explanation of the latent states and their interpretation.

Without any vaccination (a vaccination rate of 0%), the model currently predicts a tertiary wave next winter: see the blue line in Figure 2. This is in part due to the seasonal fluctuations in a subset of the model parameters; where the timing of the current secondary—and subsequent—waves rests largely upon a relaxation of social distancing, and reduced contact rates. In other words, it is societal or behavioural responses that determine the unfolding of the epidemic; rather than any loss of immunity^7^.

With an effective vaccination programme, death rates should decline more quickly with a substantial effect of vaccination evident by February. According to the upper panel in Figure 2, in mid-February a vaccination rate of 40% should reduce daily deaths from about 100 and to about 50 per day. A more aggressive vaccination programme (60% or more) could ameliorate fatalities by March 2021.

Figure 3 shows the underlying latent states generating the predictions in Figures 2. The upper panels show some outcomes from the previous figures (black dots): here, daily confirmed cases, daily deaths, and the number of patients requiring mechanical ventilation. The upper left panel shows the rates, while the upper right panel shows the cumulative totals. The remaining panels detail the fluctuations in the latent states of four factors. Each factor has two panels, showing each of the accompanying levels. For clarity, some levels have been omitted because the probabilities of being in any level—of any given factor—sum to one. Please see (Friston, Flandin et al. 2020) for a detailed explanation of the states.

The key message in this figure is an increase in the proportion of the population who are rendered immune due to vaccination (i.e., in the seronegative state). Inspection of the red lines in the right infection panel suggests that a 60% vaccination rate increases seronegative immunity from about 35% to 62%; i.e., increases immunity by 27%. This can be compared with the estimated level of 24% pre-existing (i.e., seronegative) immunity (Le Bert, Tan et al. 2020, Ng, Faulkner et al. 2020) at the beginning of the outbreak and the additional 11% of (seropositive and seronegative) immunity conferred by viral infection. This quantifies the tripartite contribution to herd immunity: (i) pre-existing immunity, (ii) exposure to the virus and (iii) effective vaccination (24%, 11% and 35%, respectively). The estimate of pre-existing immunity (24%)—i.e., 76% susceptible to attack— concurs almost exactly with the attack rate of SARS-CoV-2 in a largely unmitigated epidemic: “Correcting for cases without a detectable antibody response and antibody waning, we estimate a 66% attack rate in June, rising to 76% in October” (Buss, Prete et al. 2020).

Figure 4 shows the cumulative fatality under different rates of vaccination with current (left panel) and enhanced (right panel) levels of contact tracing. It is immediately apparent that the most effective protocol involves a combination of vaccination and enhanced contact tracing. Indeed, this combination could suppress community transmission until 2022. With current levels of contact tracing, an effective vaccination rate of 60% appears to be sufficient for suppression. This would entail vaccinating at least 25 million people by the end of next year, i.e., about 36% of the population to achieve a herd immunity of 64%.

**Figure 4:**
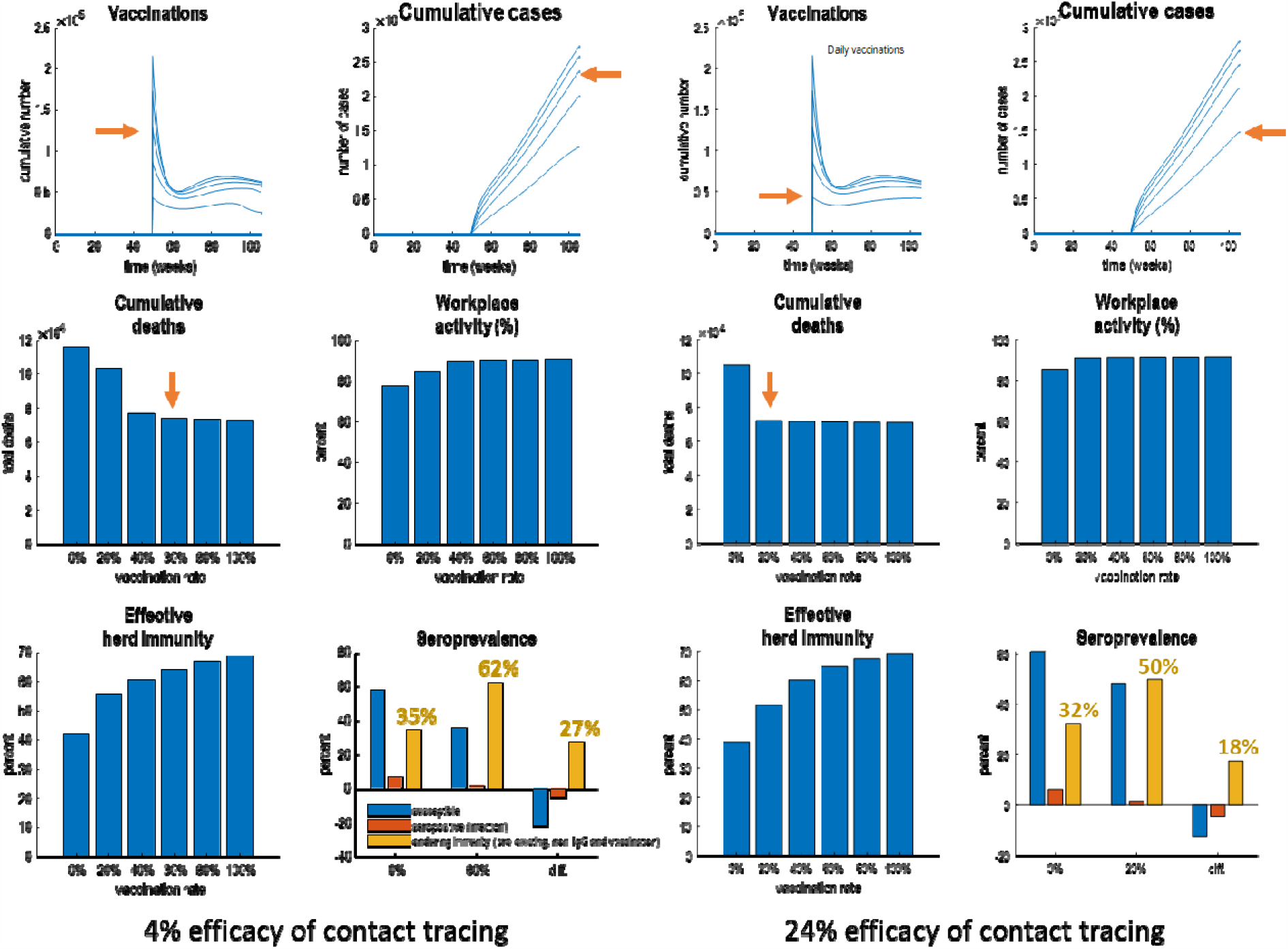
posterior predictions of mortality and economic cost under different rates of vaccination. The left panels are predictions under the current efficacy of contact tracing (about 4%), while the right-hand panels show the equivalent predictions if the efficacy of contact tracing could be increased to 24% at the time of writing (6-12-2021). In each panel, the top row describes the rate and cumulative number of (effective) vaccinations under the six rates (0% to 100% in 20% steps). These show an initial burst in vaccination rates at the inception of a vaccination programme that lasts for a couple of months that then declines exponentially to sustained rates of roughly 50,000 per day. Depending upon the rate, the total number of people vaccinated by the end of next year varies between about 12 and 26 million people. The bar charts in the subsequent three panels show the cumulative death rates, average workplace activity (based upon predictions of transport use) and effective herd immunity (the complement of the susceptible state in the DCM). Under contact tracing with an efficacy of 4%, a 60% vaccination rate would be sufficient to minimise cumulative fatalities (indicated by red arrows); inducing an effective herd immunity of about 64%. This comprises seropositive (following seroconversion due to infection) and enduring (seronegative) immunity. The 62% seronegative immunity includes pre-existing immunity, humoral immunity without detectable IgG antibodies and immunity conferred by vaccination. The contribution of vaccination can be quantified by the difference in seronegative immunity with and without vaccination; here, 27%. With enhanced contact tracing this contribution falls to 18% of a herd immunity of 50%.

This level of immunity corresponds roughly to conventional estimates of the herd immunity threshold, i.e., 1-1/*R*_0_. However, note that the proportion of people who need to be vaccinated (36%) is substantially less than the herd immunity threshold (67%) based upon simplifying assumptions. This reflects heterogeneity of exposure and susceptibility, where a substantial proportion of the population have pre-existing or acquired immunity and the effective reproduction rate depends upon mitigations—such as social distancing and self-isolation. This begs the question, could one further reduce the number of people requiring effective vaccination? The answer is yes: if the efficacy of contact tracing were increased to 24%, suppression would require a more modest vaccination rate of 20%, with 15 million people vaccinated by the end of next year. This corresponds to 22% of the population.

Figure 4 also quantifies the effect of vaccination in terms of an economic proxy; here, quantified in terms of percentage mobility based upon predictions of transport use (cars) as reported by the Department of Transport (similar predictions obtain when using Google mobility data: see Figure 1). With suppression of viral transmission, the average mobility—and implicit workplace activity— returns to about 92% of pre-COVID levels. Anecdotally, this may reflect enduring changes to social and economic behaviours; for example, working more from home and wearing masks in public places. In the model, this is mediated by the ‘long-term memory’ of exposure to the virus, encoded by the seronegative prevalence, which decays with a time constant of years (here, 2048 days).

## Conclusion

A coarse grain (countrywide) but informed quantitative (DCM) analysis of the putative impact of vaccination programmes suggest that at least 14 million people need to be vaccinated by the end of next year. In combination with pre-existing and acquired immunity, this should achieve an effective herd immunity threshold of 50%. However, this depends sensitively on enhanced contact tracing— as mandated by standard public health approaches to infectious disease control. In the absence of these public health measures, the effective herd immunity threshold increases to about 64% and would require the effective vaccination of 24 million people by the end of next year (or more, if vaccination is only partially effective).

Quantitative analyses of this sort may have important policy, health and economic implications. Currently the estimated probability of going into self-isolation when infected but pre-symptomatic is only 4%. If we expand our contact tracing effectively to support 24% of contacts in self-isolation, the modelling suggests we would reach herd immunity by vaccinating (at least) 22% of people, compared with 36% at our current level of contact isolation. This will substantially lower the potential costs of vaccination in the longer term. It will also substantially reduce the risk of recrudescence. As SARS-CoV-2 is not going away, effective tracing and isolation will ensure we can sustain a lower endemic equilibrium and reduce the risk of new outbreaks—that could have significant economic implications through local lockdowns and travel restrictions.

In principle, it should be possible to estimate the effectiveness of vaccination in a few months using the fact that the number of vaccinations times their *effectiveness* equals the number of *effective* vaccinations. In other words, one can use the DCM to fit the number of vaccinations administered by including an *effectiveness* parameter. Furthermore, with a more refined model of economic costs, it should be possible to predict both health and economic outcomes, under different vaccination programs.

### Limitations

The model described in this report is limited by its macroscopic nature. While it is sufficient to ask broad questions about the effect of one kind of intervention on another, it precludes more detailed and pragmatic scenario modelling: e.g., (Houlihan, Vora et al. 2020, Saad-Roy, Wagner et al. 2020). For example, it does not address:

- **The effects of using the vaccine to target disease prevention versus transmission**: we have simply assumed that there is a certain probability of being vaccinated when entering a clinical setting; either as a patient or a healthcare worker. A more detailed model would separately parameterise different groups—or vaccination in different settings—that may change over time.
- **Age and exposure-related effects on who is immunised:** for instance, the focus until Easter will be the elderly, who may not be contributing so much to transmission.
- **Differences in efficacy and mechanism of various vaccines:** the model assumes every vaccine is 100% effective, in preventing both infection and transmission. To model differential efficacy, one would need separate effectiveness parameters for each vaccine, which can only be estimated with appropriate data. To assess differential effects on preventing infection versus transmission would require a comparison of models in which effective vaccination increased transition from a susceptible to a seronegative state, reduced transition from infected to infectious states, or both.

As always, with dynamic causal modelling studies, the posterior predictions and assertions are conditioned entirely upon the particular model used. Although this model has been optimised using Bayesian model comparison throughout the course of the epidemic, it is just one model. As such, it is almost certainly not the best model. For example, if our assumptions about the loss of protective immunity are optimistic, perhaps through ignoring viral mutation, then the case for a more effective trace and isolation system is strengthened.

## Data Availability

All the data used is from publically available database

## Software note

The figures in this report can be reproduced using annotated (MATLAB) code available as part of the free and open-source academic software SPM (https://www.fil.ion.ucl.ac.uk/spm/), released under the terms of the GNU General Public License version 2 or later. The routines are called by a demonstration script that can be invoked by typing >> DEM_COVID_UK at the MATLAB prompt. At the time of writing, these routines are undergoing software validation in our internal source version control system—that will be released in the next public release of SPM (and via GitHub at https://github.com/spm/). In the interim, please see https://www.fil.ion.ucl.ac.uk/spm/covid-19/.

## Data availability

The data used in this technical report are available for academic research purposes from the sites listed in the legend to Figure 1.

## Acknowledgements

We are grateful to Deenan Pillay for guidance on the virological aspects of this modelling. The Wellcome Centre for Human Neuroimaging is supported by core funding from Wellcome [203147/Z/16/Z]. A.R. is funded by the Australian Research Council (Refs: DE170100128 and DP200100757).

The authors declare no conflicts of interest.

## Footnotes

1 <u>Priority groups for coronavirus (COVID-19) vaccination: advice from the JCVI, 2 December 2020 (publishing.service.gov.uk)</u>

2 <u>Report 33 - Modelling the allocation and impact of a COVID-19 vaccine | Faculty of Medicine | Imperial College London</u>

3 We obtained similar quantitative results with a shorter time constant of 1024 days.

4 <u>i-sense COVID Response Evaluation Dashboard (i-sense.org.uk)</u>

5 The prior probability over these parameters were based on the following sources and subsequently refined using Bayesian model reduction: https://royalsociety.org/-/media/policy/projects/set-c/set-covid-19-R-estimates.pdf and https://arxiv.org/abs/2006.01283.

6 Operationally, this involves comparing models in which each parameter was (i) fixed, (ii) changes as a mono-exponential function of time or (iii) a periodic function, with a duty cycle of one year. By comparing the evidence for these three models, one can then accept or reject the hypothesis that any given parameter changes with time. Clearly, this is only viable given sufficient data (e.g., the first and second surges over many months). The parameters for which there was evidence of seasonal fluctuations were the probability of developing a serious illness when symptomatic and the probability of dying from a severe illness, which decreased during the second peak.

7 <u>Microsoft Word - TR5_Second_Wave_updated_authorship.docx (ucl.ac.uk)</u>

